# Gene x Environment Interactions: Polygenic Scores and the Impact of an Early Childhood Intervention in Colombia

**DOI:** 10.1101/2025.05.11.25327374

**Authors:** Orazio Attanasio, Gabriella Conti, Pamela Jervis, Costas Meghir, Aysu Okbay

## Abstract

We evaluate impacts heterogeneity of an Early Childhood Intervention in Colombia, with respect to the Educational Attainment Polygenic Score (EA4 PGS) constructed from DNA data based on GWAS weights from a European population. We find that the EA4 PGS is predictive of several measures of child development, mother’s IQ and, to some extent, educational attainment. We also show that the impacts of the intervention are significantly greater in children with low PGS, to the point that the intervention eliminates the initial genetic disadvantage. Lastly, we find that children with high PGS attract more parental stimulation; however, the latter increases more strongly in children with low PGS.

**JEL Codes:** C21, J13, I24

## 1 Introduction

It is well established that the early years are critical to child development, with children in deprived environments accumulating deficits that are later difficult to reverse, (Heckman, 2006; Rubio-Codina, Attanasio, Meghir *et al*., 2015). Appropriate parenting interventions have been shown to compensate, at least in part, for the effects of deprivation (Grantham-McGregor, Powell, Walker *et al*., 1991; Heckman, Moon, Pinto *et al*., 2010; Grantham-McGregor, Adya, Attanasio *et al*., 2020; Schepan, Sandner, Conti *et al*., 2025) often with long-term impacts on individual cognition, earnings and health, among others (see, e.g., Gertler, Heckman, Pinto *et al*. (2014); Campbell, Conti, Heckman *et al*. (2014)). Early years interventions have therefore received a lot of attention in the policy debate on how to reduce long-term poverty and its transmission across generations, as discussed in Attanasio, Cattan, and Meghir (2022).

A frequently overlooked fact is that the effects of such interventions vary widely, with significant benefits for some groups and smaller effects for others. The well-known Jamaica study demonstrated remarkable gains in cognitive development (Grantham-McGregor, Powell, Walker *et al*., 1991), which later translated into substantial improvements in adult labor market outcomes (Gertler, Heckman, Pinto *et al*., 2014); however, the participants were malnourished children living in urban slums. In contrast, neither the Odisha intervention (Grantham-McGregor, Adya, Attanasio *et al*., 2020), that targeted the rural poor, nor the Cuttack one (Andrew, Attanasio, Augsburg *et al*., 2019), which focused on urban slum dwellers, screened for malnutrition. Their impacts were only about one third of those observed in Jamaica; nonetheless, within the Cuttack study, malnourished children experienced effects comparable to those in Jamaica, suggesting that initial disadvantage may be a key determinant of impact size. These results are consistent with established evidence that interventions appear to be more effective for children facing the greatest adversity (Conti and Heckman, 2014).

A growing body of research has highlighted the relationship between genetic background and outcomes such as education (Papageorge and Thom, 2019; Ahlskog, Beauchamp, Okbay *et al*., 2024; Biroli, Galama, von Hinke *et al*., 2025), and wealth accumulation (Barth, Papageorge, and Thom, 2017; Bartscher, Kuhn, and Schularick, 2019). These studies include genetic background not as individual genes but as a combination of genetic factors, such as those measured through polygenic scores (PGS).^1^ Educational attainment polygenic scores, for instance, have demonstrated explanatory power for educational attainment and cognitive outcomes, at least in some populations (Okbay, Wu, Wang *et al*., 2022; Chen, Kim, Lam *et al*., 2024).^2^ Rustichini, Iacono, Lee *et al*. (2023) incorporate genetic factors when modeling skill transmission across generations and find that this approach significantly alters predictions about intergenerational income elasticity (IGE).^3^ Although the underlying mechanisms and pathways remain unclear, it is widely accepted that genetic background appears to be predictive of certain phenotypes.

In this paper, we investigate whether the impacts of a randomized Early Childhood Intervention (ECI) in Colombia, documented in Attanasio, Fernández, Fitzsimons *et al*. (2014) and Attanasio, Cattan, Fitzsimons *et al*. (2020), vary depending on the child’s genetic background. This exercise is possible due to the availability of DNA from a large fraction of the children and their mothers in the study. Following the recent literature, we use a polygenic score (PGS) widely used to predict individual educational attainment and which combines a large set of genetic variations (Okbay, Wu, Wang *et al*., 2022).

An issue with the use of PGS in samples from developing countries is that most of the weights used to construct the PGS have been estimated on Western populations of European genetic ancestry, in large part due to the availability of genetic data. Therefore, before analyzing whether the intervention we consider had heterogeneous impacts depending on PGS, we must establish that such a PGS is predictive of child development in the population we study. This evidence is interesting in its own right.

Our paper is one of the first to consider how the impact of an early education intervention in an LMIC context is moderated by polygenic scores that generate possible heterogeneous impacts based on genetic background. A study close to ours is Biroli, Galama, von Hinke *et al*. (2025), which considers whether a natural experiment that changes mandatory schooling attenuates the disadvantage associated with genetic background and, more generally, discusses the issues involved in the use of genetic data and the interactions between genetic background and environmental factors.^4^ Houmark, Ronda, and Rosholm (2024) study the association of genetic background with the process of human development, looking at a structural model that includes child development, parental investment, and genetic variables.

Therefore, we innovate in two dimensions: we provide some validation of the specific PGS we use, in a context where genetic information is scarce, and we show how a randomized intervention interacts with this PGS to produce heterogeneous impacts on early childhood development.

First, we show that the PGS we use is predictive of child cognitive development both at baseline and at the first follow-up of the randomized controlled trial (RCT) we study, when the children were about 30 months old. We also show that caregivers’ PGS is predictive of their own performance in the Raven test, a measure of intelligence, and, to an extent, correlated with their educational attainment. Interestingly, child PGS is also predictive of parental investment, in agreement with the evidence presented by Wertz, Moffitt, Agnew-Blais *et al*. (2020).

Child PGS’s are obviously correlated with parental PGS’s. This correlation makes it difficult to interpret the relationship between the child’s PGS and the developmental outcomes. As already observed in the literature (Wertz, Moffitt, Agnew-Blais *et al*., 2020), it is possible that parental and child PGS’s are correlated with observed and unobserved investments in children, including social influences reflected in parental phenotypes and correlated with child PGS. These difficulties can severely alter the interpretation of the role of child PGS.

Second, we find that the impacts documented in Attanasio, Fernández, Fitzsimons *et al*. (2014) and Attanasio, Cattan, Fitzsimons *et al*. (2020) are heterogeneous by the PGS of children. In particular, we find that the impact of the intervention is much higher in children with relatively low PGS, which points to lower genetic propensity to acquire education. The difference is such that the intervention seems to eliminate the “genetic disadvantage” experienced by children with low PGS. This result shows an important gene-environment interaction and is one of the first to be documented within an RCT designed to evaluate a specific intervention.

Interestingly, we obtain a similar result for parental investment. Attanasio, Cattan, Fitzsimons *et al*. (2020) show that the intervention had a strong and positive impact on some components of parental investment and argue that most of the impact is mediated by increased investment. Here we find not only that high PGS children receive higher parental investments, but also that the impact of the intervention on parental investment is particularly high in children with a *low* PGS.

The remainder of the paper is organized as follows. In Section 2, we describe the RCT performed in Colombia and the data collected to evaluate it, including DNA material. In Section 2.3, we discuss the PGS used in this context. In Section 3, we show that such a PGS is predictive of both children’s and caregivers’ outcomes. Finally, in Section 4, we report the results of the impact heterogeneity by PGS. Section 5 concludes the paper.

## 2 Data and Context

The data we use in this article was collected in connection with a randomized controlled trial that was carried out in the central region of Colombia in 2008-2009. In this section, we briefly describe the intervention and its impacts, as well as the context within which the DNA data we use was collected.

### 2.1 An early years intervention in Colombia and its impacts

In 2010, a RCT was used to evaluate: (i) an ECD parenting intervention focused on child stimulation and inspired by the Reach Up program (Jervis, Coore-Hall, Pitchik *et al*., 2023), (ii) a nutrition intervention (which consisted of the delivery of nutritional supplements), and (iii) their interaction. 96 towns, with a population between 2000 and 42000 in eight departments in Colombia, were randomly allocated to four groups: a control sample, a stimulation sample, a nutrition sample and a sample that received both interventions. The intervention, which lasted 18 months, targeted families with children aged 12 to 24 months at its start and who were beneficiaries of a Conditional Cash Transfer (CCT) program in Colombia. The intervention was carried out by local women, who were beneficiaries of the same CCT and who had been elected as representatives of groups of beneficiaries. These women, called *Madres Lĺderes* (ML), were hired and trained (in treatment towns) to deliver the appropriate intervention. During the duration of the intervention, they were mentored and advised by the same professionals who trained them at the start of the study.

The 1440 children in the sample were administered a variety of developmental tests at baseline and at the end of the study, approximately 18 months later. Various anthropometric measures were also obtained, including height, weight, and anemia. Their families were administered a rich survey that included standard socioeconomic and demographic variables as well as parent-reported measures of child development.

The impacts of the two interventions and their interaction were reported in Attanasio, Fernández, Fitzsimons *et al*. (2014). The first noteworthy result is that the nutrition intervention, which included the delivery of micro nutrients, did not have any effect, somewhat surprisingly, as the prevalence of anemia at baseline was high in the sample. However, the stimulation intervention, both alone and in combination with the nutrition intervention, was shown to have sizable and statistically significant effects on child development, which was measured using the Bayley Scales of Infant Development (BSID), and other scales. For example, the cognitive scale of the BSID improved by 0.26 of a standard deviation, with a p-value of 0.002, while the receptive language scale improved by 0.22 of a standard deviation (p-value =0.032).^5^

In a subsequent paper, Attanasio, Cattan, Fitzsimons *et al*. (2020) show that the stimulation intervention increased parental investment in children, both in terms of time and materials. Furthermore, the mediation analysis performed in that paper shows that the impact of the stimulation intervention can be almost completely explained by the observed increase in parental investment.

Several years after the end of the intervention, in 2013, a second follow-up was collected for this sample. In this survey, in addition to information on child development, parental investment, and other standard information, DNA material for both the children in the evaluation sample and their principal caregiver was also collected, which we discuss in the following section. The evidence from the second follow-up indicated that the impacts observed at the end of the interventions on child development had waned completely, as discussed in Andrew, Attanasio, Fitzsimons *et al*. (2018). Similarly, child investment also decreased, which may have contributed to the fading out of the impacts.

Given this evidence, in the analysis that follows, we focus on the child development outcomes at the first follow-up. In particular, we first consider whether child development outcomes are predicted by genetic background. We also consider whether the PGS’s available to us are predictive of parental investment and of mothers’^6^ outcomes. As discussed in the Introduction, these results are important in their own right. Having shown the relevance of the DNA data, we consider the possibility that the impacts at the end of the intervention are heterogeneous depending on children’s genetic background, that is, we study the interaction between the educational attainment polygenic score and an environmental factor.

### 2.2 DNA procedure and data

DNA material was obtained through buccal swabs (gum mucosa), which were collected from the target children and their principal caregivers (PC) during the second follow-up phase of the study, with the aim of investigating whether genetic factors influenced the impact of the intervention. DNA obtained from buccal cells is comparable to the material obtained by blood collection.

Qualified staff employed by the data collection company were trained to assist in the process. Buccal swabs were collected exclusively from households that consented to participate. The procedure was carried out in a health center or community hall within each of the sample villages after obtaining consent from the participants. We provide additional details on the DNA data collection and the consent procedure in the Appendix.

DNA material was obtained from 986 caregivers who consented (a consent rate of 78%, based on a total follow-up sample of 1,257 observations) and 1,080 target children (a consent rate of 86%). In particular, we obtained 983 complete caregiver-child pairs; for 97 children, PC DNA was not collected; finally, we had 3 children for whom we collected PC DNA but not child DNA. This collection rate compares well with large studies such as the Fragile Families and Child Wellbeing Study (FFCWS) and the UK Millennium Cohort Study (MCS), where just over 83% of the eligible mothers provided a saliva sample (see Ware Erin (2021); Fitzsimons, Moulton, Hughes *et al*. (2022)).

As mentioned above, the DNA samples were held in the utmost confidentiality and were not linked directly with the household ID or any personal identifier. Unfortunately, difficulties in field work, which we discuss in detail in the Appendix Section A.2, made it impossible to link some of the DNA material to the survey data and to the children’s results. This led to the loss of sample units. Below we present evidence that the observations are missing at random.

DNA extraction was performed at the Genome Center of Queen Mary University of London; samples were then assayed with the Illumina Global Screening Array (GSA) at the HuGe-F facility of Erasmus University Rotterdam (mothers’ sample) and at King’s College London NIHR Maudsley Biomedical Research Centre (BRC) (children’s sample). Further details on the genotyping are provided in the Appendix. Due to duplicates, poor quality performance, low stock volume, or failed Qubit measurements, the sample size assayed with Illumina GSA was reduced to a total sample of 1,362 individuals, consisting of 656 (out of 986) caregivers, and 706 (out of 1,080) target children, yielding 522 (out of 983) caregiver-child pairs.

In addition, we performed extended quality control to check: i) that the phenotypic gender is confirmed by the extracted DNA, ii) family relationship (most principal caregivers are the biological mothers) and iii) ancestry (most samples have Colombian, Mexican, or Puerto-Rican ancestry, as expected). Given all this, the data of the target children sample we use is restricted to 528 observations (out of 706). In the analysis we report below, we also exclude 2 extreme outliers, defined as those with a PGS more than 4 SDs distant from the mean.

In the Appendix, Table A1 presents balance tests for the sample in which we have linked child genetic information, as well as for the sample in which we have DNA data for the primary caregivers. Reassuringly, the characteristics of the DNA sample are well balanced, with no significant differences between treatment and control samples.

Given the limited size of the sample for which genetic data is available, we further test whether DNA data are missing at random. The outcome variable is a cognitive score factor constructed from three Bayley subscales: cognition, receptive language, and expressive language.^7^ In Table 1, we regress the outcome of the intervention on treatment status and a set of controls, including child age and gender, a quadratic in mother’s age, interviewer’s dummies, and baseline values for: Bayleys and its square, mother education dummies, a poverty index, indicators for anemia and stunting, and mother depression. Column (1) reports the results on the entire sample. In column (2), we add a missing DNA variable: this is not significant and does not change the estimated impact much. Finally, in column (3), we estimate the impact only on the subsample where the DNA is not missing. The impact remains almost unchanged, albeit with a higher standard error reflecting the loss in the sample.^2^ The conclusion is that the DNA data is missing at random, and that the cost of this loss is not bias, but power to detect effects.

**Table 1.** DNA data missing at random: Child Cognitive Development.

|  | Complete sample<br>(1) | Complete sample<br>(2) | Non missing DNA<br>(3) |
| --- | --- | --- | --- |
| treat | 0.164***<br>(0.057) | 0.169*<br>(0.089) | 0.146*<br>(0.086) |
| Missing DNA |  | 0.037<br>(0.060) |  |
| treat on<br>missing DNA |  | -0.008<br>(0.105) |  |
| Observations | 1144 | 1144 | 437 |
| $R^2$ | 0.315 | 0.315 | 0.323 |

This Table displays coefficients from linear regressions of follow up Bayley’s cognitive index on an indicator of whether the child was treated or not. In column 2, the regression also includes a dummy that indicates the observations for which DNA information is missing and the interaction of this dummy with treatment status. In column 3, estimation of treatment effect is limited to the sample with non-missing DNA information. The set of controls included in all columns are child age and gender, a quadratic in mother’s age, interviewer’s dummies, and baseline values for: Bayleys and its square, mother education dummies, a poverty index, indicators for anemia and stunting, and mother depression. Standard errors (in parenthesis) are clustered at the village level. \* $p < 0.10$ , \*\* $p < 0.05$ , \*\*\* $p < 0.01$ . <sup>a</sup> The RW p-values are adjusted for 2 hypotheses in column 2, for 3 hypotheses in column 3 and 4 hypotheses in column 4.

### 2.3 The Educational Attainment Polygenic Score

Polygenic scores (PGS) are genetic predictors that aggregate the estimated effects of many SNPs on a phenotype *y*. A typical PGS is computed from SNP information as follows.

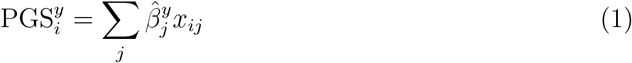

for all individuals *i*. In equation (1) *x*_*ij*_ is *i*’s genotype at SNP *j* taking the values 0, 1, or 2 depending on whether the individual has inherited 0, 1, or 2 copies of the allele in question. 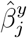 is the strength of the association of that allele with the trait in question, which in our case is educational achievement. These parameters are estimated in very large samples, typically from Western populations.

Although individual genetic variants have tiny effects on complex phenotypes such as social and behavioral outcomes, PGSs are much more predictive. The PGS we use in this study is a genetic predictor for educational attainment. It was constructed using the estimated effects of SNPs from a genome-wide association study (GWAS) of educational attainment carried out on a sample of *∼* 3 million individuals of European genetic ancestry (Okbay, Wu, Wang *et al*., 2022). This PGS is reported to explain 12.9% and 17% of the variance in educational attainment for individuals of European genetic ancestries in the Health and Retirement Study (HRS) and the National Longitudinal Study of Adolescent to Adult Health (Add Health) samples. We report details on the computation of the PGS we use in the Appendix.

PGS have been demonstrated to have poor predictive power when constructed using SNP effects estimated in GWAS discovery samples of a genetic ancestry that is different from the validation sample’s. For example, Okbay, Wu, Wang *et al*. (2022) estimate the predictive power of the educational attainment PGS to be 1.3% and 2.3% for individuals of African genetic ancestry in HRS and Add Health, respectively, representing an attenuation of 89% and 85%. Various factors have been shown to contribute to this drop in predictive power, including but not limited to differences between populations in causal variant effect sizes, in alleles frequencies and in the correlation between causal variants and SNPs included in the GWAS, as well as gene*×*environment interactions. In general, the portability of PGS decreases with increased genetic differentiation between ancestries (Wang, Guo, Ni *et al*., 2020).

The Colombian population is known to be an admixture of genetic ancestries from the Americas, Europe, and Africa, with percentages depending on the region (Ossa, Aquino, Pereira *et al*., 2016). As such, we expect the decrease in the predictive power of the PGS in our sample to be lower than that observed in Okbay, Wu, Wang *et al*. (2022) for individuals of African genetic ancestry, provided that the dominant factor contributing to the loss is genetic differences rather than gene*×*environment interactions. The extent to which the PGS we consider is predictive of developmental outcomes in our sample is therefore an interesting question.

## 3 Is the PGS Predictive of Maternal and Child Outcomes?

Having collected DNA data and calculated an educational attainment polygenic score, we can evaluate its predictive validity in the current context. In particular, we check whether the polygenic scores we calculate are predictive of a set of different outcomes. Such an exercise is interesting in its own right, given the paucity of data from developing countries and non-Western populations. At the same time, it will be the first step in establishing whether the impacts of the intervention implemented in the population considered are heterogeneous by values of the polygenic scores. To mitigate any measurement error and to make the interpretation of the estimated coefficients easier, we decided to use a binary indicator throughout for PGS above/below our own sample median.^9^

Before showing the extent to which the PGS is predictive of child development, we should stress that these correlations are not causal. The PGS can be correlated with parental genotypes and phenotypes, as well as with other socioeconomic variables that we do not observe and that could be the main causal factors driving child development.

### Predicting Child Developmental Outcomes with an Education PGS

We start by considering the predictability of the cognitive development of children by PGS. These are reported in the first four columns of Table 2. We used the subsample of our survey in which the DNA information was successfully matched, allowing the calculation of the educational attainment PGS. As we discussed above, DNA is missing at random, so the sample we are using is representative. For this sub-sample, we construct a cognitive measure that combines three subscales of the Bayley Scales of Infant Development III (namely, the cognition, expressive, and receptive language subscales). These measures were collected both at baseline, when the children were about one year old, and 18 months later, at the first follow-up of the study, which we will use for the main analysis of this paper.

**Table 2.** Predictability of child and mother (PC) outcomes by their respective PGS.

|  | Child Cognitive Domain - Bayley |  |  |  | Maternal Cognition and Education |  |  |  |
| --- | --- | --- | --- | --- | --- | --- | --- | --- |
|  | (1)<br>Baseline | (2)<br>Follow-up | (3)<br>Baseline | (4)<br>Follow-up | (5)<br>Raven | (6)<br>Years of Ed | (7)<br>Raven | (8)<br>Years of Ed |
| PGS<sup>a</sup> ≤ median | -0.289***<br>(0.095)<br>[0.002] | -0.324***<br>(0.101)<br>[0.002] | -0.175**<br>(0.087)<br>[0.081] | -0.193**<br>(0.089)<br>[0.081] | -2.551***<br>(0.933)<br>[0.027] | -0.805*<br>(0.435)<br>[0.048] | -1.884**<br>(0.925)<br>[0.103] | -0.498<br>(0.435)<br>[0.230] |
| Controls | Basic | Basic | Full | Full | Basic | Basic | Full | Full |
| Observations | 491 | 479 | 450 | 437 | 323 | 312 | 301 | 299 |
| R<sup>2</sup> | 0.081 | 0.125 | 0.162 | 0.356 | 0.103 | 0.091 | 0.178 | 0.184 |

<sup>a</sup>In **Columns 1-4** PGS is that of the child. In **Columns 5-8** PGS is that of the mother. In the first two columns, a basic set of controls includes the first 20 principal components of the child's SNPs as well as interviewers' dummies and child gender and age in months. In column 3 and 4, the set of controls also includes indicators for stunting and anemia at baseline as well as a PC's age and its square, an indicator for depression at baseline, a poverty index. The RW p-values are adjusted for 2 hypotheses in columns 1 and 2 and for 2 hypotheses in columns 3 and 4 reported in square brackets. In **Columns 5-8** Basic controls include interviewer dummies and 20 principal components of the PGS's SNPs. Full control also include PC's age and a depression index as well as a poverty index. The RW p-values are adjusted for 2 hypotheses in column 5 and 7 and 2 hypotheses in column 6 and 8 in square brackets. Standard errors (in parenthesis) are clustered at the village level. \* $p < 0.10$ , \*\* $p < 0.05$ , \*\*\* $p < 0.01$ . <sup>a</sup> Standard errors (in parenthesis) are clustered at the community level.

In addition to the PGS variable, we add interviewer dummies and controls for child gender and age in months, and strata controls. We also control for child’s indicators of stunting and anemia at baseline, as well as for the Principal Caregiver’s age and its square, education, depression, and an index of poverty. Furthermore, as is common practice in the literature on PGS, we also include the first 20 principal components of genomic data to control for confounding due to population stratification (Price, Patterson, Plenge *et al*., 2006). This set of controls will also be used in the analysis of the interaction between treatment effects and genetic background.

The results indicate that the PGS can predict the baseline Bayley cognitive score. In particular, at baseline and follow-up, children with a PGS below the median have measures of cognitive development −0.29 and −0.32 standard deviations lower, respectively, than children with a PGS above the median when we use a basic set of controls, including 20 principal components of the PGS SNPs, interviewer dummies and child age and gender. These associations are strongly significant with a Romano Wolf-corrected p-value of 0.002. When we add a richer set of controls, including Principal Caregiver age, education and depression, as well as an indicator of stunting and one of poverty, the association is much lower, with coefficients of 0.18 and 0.19 at baseline and follow-up, respectively. These coefficients have RW adjusted p-values of 0.08.^10^ Thus, there is some evidence that PGS predicts lower child performance.

### Predicting Primary Caregiver Education Attainment

We now turn to the predictability of adult outcomes among the principal caregivers (PC) in our sample with respect to their own educational PGS^11^. The results are reported in columns 5-8 of Table 2. We consider two outcomes, the Raven score obtained from the principal caregiver, and their educational attainment, measured by the number of years of formal schooling. We report two sets of results, one with only a basic set of controls, which include interviewers’ dummies and the first 20 principal components of the PC genomic data, and another one with an extended set, which also includes PC’s age and depression index at baseline and a poverty index. The p-values for the significance of the PGS coefficients are adjusted for multiple hypotheses testing (considering the Raven test and education).

When considering only the basic set of controls, the coefficient on the low PGS dummy is negative and strongly significant for the Raven tests. Under conventional single-hypothesis standards, the p-value for the predictability of the Raven when we include only basic controls is 0.008. For educational attainment, the coefficient is also negative with an adjusted RW p-value of 0.048. When controlling for some additional variables, such as age, depression, and a poverty index, the size of the coefficient decreases, and the RW adjusted p-values for the two coefficients are 0.103 and 0.230. Considering our relatively small sample, we interpret the results on the correlation between Raven scores and PGS as quite strong evidence of predictability. However, in this sample, we find little evidence that it predicts educational attainment, possibly reflecting the limited variability of education in the sample.

### Investments and the PGS

Finally, we examine whether parental investments in children are predicted by child PGS. We include the same controls as for the regressions in Table 2. The investment measures for materials are composites of books and developmentally appropriate toys found in the household, while time investments are a composite of time spent with the child in developmentally relevant activities. These measures were collected during the baseline and follow-up interviews and are described in detail in Attanasio, Cattan, Fitzsimons *et al*. (2020). Observing the predictability of these investments based on the child PGS points to possible mechanisms through which child PGS may affect final outcomes.

Table 3 shows the results. In all cases, the estimates point to lower investments for children with low PGS, which would be consistent with their lower level of achievement, shown in Table 2. However, the effects become considerably smaller and mostly insignificant once we include the full set of controls, both for material and time. The implication is that observable characteristics of mothers and children that are correlated with PGS also drive investments.

**Table 3.** Parent Investment at Baseline.

|  | Material Investment |  | Time Investment |  |
| --- | --- | --- | --- | --- |
|  | (1) | (2) | (3) | (4) |
| Child PGS<br>≤ median | -0.204**<br>(0.086)<br>[0.043] | -0.128<br>(0.084)<br>[0.226] | -0.173***<br>(0.063)<br>[0.050] | -0.110*<br>(0.063)<br>[0.266] |
| Controls | basic | full | basic | full |
| Observations | 465 | 453 | 465 | 453 |
| $R^2$ | 0.136 | 0.227 | 0.129 | 0.221 |

Dependent variables: Columns 1-2 Material Investment at baseline; Columns 3-4 Time Investment at baseline. A basic set of controls, included in columns 1 and 3, are child age and gender, a quadratic in mother’s age, interviewer’s dummies and the first 20 principal components of the PGS’ SNPs. In columns 2 and 4, the set of controls also includes indicators for mother’s depression at baseline, education dummies, a poverty index and an indicator for stunting at baseline. Standard errors (in parenthesis) are clustered at the village level. \* $p < 0.10$ , \*\* $p < 0.05$ , \*\*\* $p < 0.01$ . <sup>a</sup> The RW p-values, reported in square brackets are adjusted for 2 hypotheses in column 1 and 3 and 2 hypotheses in column 2 and 4.

## 4 The Polygenic Score and The Intervention Impacts

In this Section, we examine the possibility that the intervention had different impacts on children with different polygenic scores, which we have shown to be predictive of several developmental outcomes. As mentioned in Section 2.2, the outcome variable we consider is the cognitive score factor constructed from the three Bayley subscales. Similar results are obtained with the individual subscales (cognition and receptive language) for which we record a positive impact.

### Heterogeneous Intervention Impacts on Child Development

Our main results on the differential impacts of the interventions are presented in Table 4. In column (1), for ease of presentation, we report again the estimates without any DNA indicators, as in Table 1. In column (2), we report estimates of the impacts of the intervention for children with a value of the PGS above and below the median. Since we are testing two hypotheses (impact of the intervention for low and high PGS) we present Romano and Wolf (2005) stepdown pvalues (RW) that adjust for multiple hypotheses testing.

**Table 4.** Treatment impact heterogeneity on Bayley cognitive index by PGS and education.

|  | (1) | (2) | (3) | (4) |
| --- | --- | --- | --- | --- |
| treatment | 0.146*<br>(0.086) |  | 0.424***<br>(0.121) [0.003] |  |
| treatment × low PGS | | 0.287**<br>(0.121) [0.043] | | 0.538***<br>(0.159) [0.002] |
| treatment × high PGS | | -0.041<br>(0.119) [0.720] | | 0.255*<br>(0.137) [0.079] |
| treatment × complete primary | | | -0.320*<br>(0.160) [0.050] | -0.343**<br>(0.166) [0.028] |
| treatment × more than primary | | | -0.512***<br>(0.175) [0.007] | -0.487***<br>(0.170) [0.01] |
| Child PGS ≤ median | | -0.299***<br>(0.108) | | -0.279**<br>(0.107) |
| Observations | 437 | 437 | 443 | 437 |
| $R^2$ | 0.323 | 0.380 | 0.338 | 0.393 |
| p. val. treatment × low PGS=<br>treatment × high PGS | | 0.051 | | 0.084 |

Dependent variable Bayley's cognitive index. A set of controls included in all columns are child age and gender, a quadratic in mother's age, interviewer's dummies, and baseline values for: Bayleys and its square, mother education dummies, a poverty index, indicators for anemia and stunting, and mother depression. The regression which include the child PGS also include the first 20 principal components of the PGS' SNPs. Standard errors (in parenthesis) are clustered at the village level. \* $p < 0.10$ , \*\* $p < 0.05$ , \*\*\* $p < 0.01$ . <sup>a</sup> The RW p-values, reported in square brackets, are based on 2 hypotheses in column 2, 3 hypotheses in column 3, and 4 hypotheses in column 4.

According to these results, the intervention completely compensates for low PGS, which is associated with disadvantage, as shown in Table 2. The size of this compensating effect can be seen in column (2) by comparing the coefficient of the low PGS dummy (−0.299) to that interacted with treatment (0.287, RW p-value 0.043). The intervention appeared to have no impact for children with high PGS (−0.041, RW p-value 0.7). The difference between the impacts is significant, implying that the intervention did little for children with higher PGS, an issue that is interesting and troubling at the same time, since these children are not well off by any definition.^12^

In columns (3) and (4), we explore heterogeneity with respect to primary caregiver education (PC) and by both PGS and education. Our definition of education (less than primary, complete primary, and more than primary) splits our sample roughly into three equal parts. Column (3) implies that the impact of the intervention can be accounted for by children whose mothers completed less than primary education. Again, the impact seems to be greater for the most disadvantaged children.

In column (4), we consider together heterogeneity by the education of the PC and by PGS. Our point estimates indicated that the greatest impact of the intervention is for children with low PGS whose mothers have education less than primary; next is the group with high PGS and mothers with this lowest education. These results tell a more complete story: the intervention improved the outcomes of children whose mothers have the lowest level of education, with the effect being greater for those in this group who have a low PGS (p-value for the difference 0.084). It appears that part of the heterogeneity attributed to the PGS can be explained by mother’s education, with the intervention having a stronger impact on the children of mothers with the lowest education.

### Heterogeneous Intervention Impacts on Parental Investment

To interpret these results, we now turn to the parental investment behavior, which was shown to be the key mediator in this intervention by Attanasio, Cattan, Fitzsimons *et al*. (2020). Table 5 presents the results for time investments (columns 1-4) and material investments (columns 5-8), mirroring the specifications we estimate for child development in Table 4.

**Table 5.** Time and Material Investment.

|  | Time Investment |  |  |  | Material Investment |  |  |  |
| --- | --- | --- | --- | --- | --- | --- | --- | --- |
|  | (1) | (2) | (3) | (4) | (5) | (6) | (7) | (8) |
| treatment | 0.199**<br>(0.083) |  | 0.330***<br>(0.123)<br>[0.005] |  | 0.117<br>(0.087) |  | 0.218**<br>(0.106)<br>[0.059] |  |
| treatment ×<br>low PGS |  | 0.325***<br>(0.105)<br>[0.006] |  | 0.457***<br>(0.139)<br>[0.001] |  | 0.128<br>(0.115)<br>[0.273] |  | 0.199<br>(0.137)<br>[0.164] |
| treatment ×<br>high PGS |  | 0.082<br>(0.126)<br>[0.501] |  | 0.248<br>(0.150)<br>[0.109] |  | 0.087<br>(0.122)<br>[0.473] |  | 0.182<br>(0.135)<br>[0.197] |
| treatment ×<br>primary |  |  | -0.258*<br>(0.146)<br>[0.051] | -0.264<br>(0.163)<br>[0.128] |  |  | -0.079<br>(0.140)<br>[0.590] | -0.012<br>(0.152)<br>[0.960] |
| treatment ×<br>≥ primary |  |  | -0.167<br>(0.167)<br>[0.180] | -0.199<br>(0.162)<br>[0.2216] |  |  | -0.220<br>(0.143)<br>[0.140] | -0.227<br>(0.146)<br>[0.134] |
| Child PGS<br>≤ median |  | -0.078<br>(0.116) |  | -0.066<br>(0.114) |  | -0.131<br>(0.108) |  | -0.126<br>(0.106) |
| Observations | 451 | 451 | 457 | 451 | 451 | 451 | 457 | 451 |
| $R^2$ | 0.266 | 0.316 | 0.262 | 0.313 | 0.300 | 0.351 | 0.303 | 0.354 |
| p.val.treat. × lowPGS<br>=treat × high PGS |  | 0.137 |  | 0.825 |  | 0.307 |  | 0.913 |

Dependent Variable columns 1-4 Time Investment and Columns 5-8 Material Investment. A set of controls included in all columns are child age and gender, a quadratic in mother's age, interviewer's dummies, and baseline values for: Bayleys, time investment, mother education dummies, a poverty index, indicators for anemia and stunting, and mother depression. The regressions which include the child PGS also include the first 20 principal components of the PGS' SNPs. Standard errors (in parenthesis) are clustered at the village level. \* $p < 0.10$ , \*\* $p < 0.05$ , \*\*\* $p < 0.01$ . <sup>a</sup> The RW p-values are in square brackets and are adjusted for 2 hypotheses in column 2, for 3 hypotheses in column 3 and 4 hypotheses in column 4.

Consistent with the results for the entire sample, treatment increased both time (col 1) and material (col 5) investments, although the latter is not significant in our smaller sample. When we interact treatment with low/high PGS we find that time investments increase much more for children with low PGS than for those with high PGS, and the difference is significant at the 10% level. The pattern is similar for material investments, but unfortunately the effects are too noisy relative to their size to draw stronger conclusions.

Similarly to the evidence on the impacts of intervention on child development, in columns (3) and (7), we also explore heterogeneity by mother’s education. Consistent with the results on child development, there is some suggestive evidence that the increase in investment came from mothers with the lowest education for both time and material investments. The impact on time investment is strongly significant for mothers with less than primary education. However, although the point estimates indicate smaller impacts for mothers with higher education, these impacts are not statistically significant.

In columns (4) and (8), we include all interactions (education and low/high PGS). For time investment, we find that most of the investment increase came from mothers with low PGS children. For materials, the picture is again less clear.

### Summary

The results we presented show that the intervention has a strong impact for children with low PGS and particularly for those with low-educated mothers. We do not have the power to distinguish whether the PGS or the education moderator are the main source of heterogeneity. However, these results suggest that the intervention closed the gap between the least and most deprived, without changing much for the former.

When we consider investment behavior, a possible explanation emerges: the intervention mainly increased time investments for the least better off and not as much for the rest. In Table 3, we show that children with lower PGS seem to attract less parental investment. Therefore, on the one hand, these results indicate that managing to increase parental investments is key to the success of such an intervention and that the least advantaged can benefit substantially (at least in the short run). On the other hand, we need to ask the question as to why mothers who are better educated or have children who are less disadvantaged, as implied by the high PGS, do not respond as much to the intervention that promotes engagement with children.

## 5 Conclusions

In the context of a randomized early childhood intervention in Colombia that was shown to have significant short-term results, we examine the heterogeneity of impacts by a genetic marker, the educational polygenic score (PGS), and by the education of the mother.

The use of PGS in this population and for this purpose is novel. We show that child PGS is predictive of child outcomes. The mother’s own PGS is less predictive of her IQ and education, once we control for observable characteristics. We then show that the intervention mainly affected children with low PGS and those whose mothers had the lowest education. Finally, we show that the intervention increase parental investment more for children with a relatively low PGS (who also, in the control group, attract less investment). In our sample, we do not find large and significant differences in the impacts on investment by mothers’ education.

There are two broad conclusions from these results. First, the educational PGS we use in this population is predictive of child development and, therefore, summarizes characteristics that indicate disadvantage. Second, the Colombian intervention we have studied mainly affected the least-disadvantaged children, based on their own PGS or based on the mother’s education.

The results we obtain in this paper point to an interesting research agenda that would be made possible by the availability of genetic data in studies of early childhood interventions. Richer data, including large samples where DNA observations of *both* parents as well as the child were available, would be important to control for confounding factors arising from the correlation of parental DNA with both parental phenotypes and child genetics. This would also allow us to better understand how the genetic background relates to parental behavior toward children (such as investments in their upbringing) and to better explore the role of gene-environment interactions in mediating the impact of interventions. Recognizing heterogeneity by genetic background can in principle allow us to design improved interventions in a way that improves the well-being of all children.

Finally, it is important to realize that all children in our sample are deeply deprived, whether they have a high or a low PGS. While it is comforting to know that the intervention compensated for the deprivation associated with a low PGS, having the ability to improve the results for all, including those with a high PGS is of fundamental importance.

## Data Availability

All data produced in the present study are available upon reasonable request to the PI (Orazio Attanasio) upon publication.

## Appendix A

## A.1 DNA data collection details

The DNA data collection was started requiring the participants to provide consent. In particular, the consent form requested permission to collect buccal cell samples from target children and/or their caregivers. Consent could be provided by caregiver, the caregiver on behalf of the child, or for both him/herself and the target child. To minimize nonparticipation, caregivers were asked to collect their own and/or their child’s buccal cell samples. The consent form outlined the procedure for collecting the buccal mucosa, the associated risks and benefits, and emphasized that the procedure was painless and completely safe. It also detailed the measures taken to protect participants’ confidentiality and security.

Upon signing the consent form, the staff explained the buccal cell collection procedure to the mother using a set of five posters designed to clearly illustrate the instructions. In particular, each poster stressed the following points:

- The procedure takes no more than a few minutes per subject.
- To minimize external contamination, both the staff and the caregiver wore protective gloves and face masks when handling materials.
- The caregiver received a glass of water to rinse her mouth and remove any food residue (staff provided water and glass).
- The caregiver was asked to massage his/her cheeks with his/her fingers to stimulate saliva production. The caregiver then used 2 to 3 cotton swabs to wipe the inside of her cheeks and tongue for 20 seconds each, placing each swab in a tube containing a solution that will break cells and extract DNA (using the Gentra Puregene Buccal Cell Kit (400) Cat No 158867, sold by qiagen). A unique code was recorded both on the sample tube and in the questionnaire.
- The caregiver then repeated the same procedure for the target child, using a new tube for the child’s sample.

Following a recommendation from the Research and Ethics Committee, each sample was identified in the database using a unique code, distinct from the participant’s ID used in the rest of the survey, and without any personal identifiers. This measure implied that participants’ data could not be linked to their identities, making individual results untraceable.^13^

## A.2 DNA Collection and linkage to Survey

In collecting DNA material, we strove to maintain confidentiality. Unfortunately, the data collection company interpreted the commitment to confidentiality too strictly and discarded the personal identifier before matching the DNA data with the household survey and the children’s developmental measures, despite the fact that IRB approval was obtained for this link. Eventually, linking the DNA data to the survey was made possible by the fact that the interviewers collecting the DNA were instructed to label the tubes sequentially in each village. Furthermore, the staff recorded location data and followed a certain order, which was preserved on their field work sheets. Since there are on average only 15 households per locality, we were able to reproduce the interview patterns from the field work sheets to associate the unique code of the DNA tube with the unique individual identifier of the survey data set for 93% of DNA observations. However, labeling instructions were not followed everywhere. For 145 tubes it was not possible to link the unique code of the DNA tube with the unique individual identifier of the survey data set. As a consequence these data are not used in this paper.

## A.3 Genotyping

The analysis of the DNA material included genotyping and epigenotyping assessments to evaluate the role of genetic factors in response to the intervention. The DNA was assayed using the Illumina Global Screening Array (GSA; HuGe-F at Erasmus and King’s College London lab), suited for multi-ethnic samples. It also included extended quality control and imputation.

## A.4 Details on PGS computation

GWAS are performed by running regressions of the phenotype on each of millions of SNPs separately, controlling for some basic covariates. Because of this, estimated SNP effects capture the effect of the predictor SNP as well as those around it that are correlated with it. The correlation between proximal SNPs, called “linkage disequilibrium (LD)” arises because the probability of co-inheritance is inversely proportional to the distance between two SNPs. Therefore, including all SNPs in a PGS results in double-counting of effects. To address this issue, we followed the same procedure as Okbay, Wu, Wang *et al*. (2022) and adopted a Bayesian methodology, called SBayesR (Lloyd-Jones, Zeng, Sidorenko *et al*., 2019), implemented in the software package GCTB (Zeng, De Vlaming, Wu *et al*., 2018). SBayesR imposes a flexible finite mixture of normal distributions on the SNP effects. To estimate the posterior mean SNP effects adjusted for LD, it requires a reference data set containing correlation estimates between SNPs. For this purpose, we used the LD matrix for the 2,865,810 pruned common variants from the full UKB European-genetic-ancestry (*N ≈* 450, 000) data set from Lloyd-Jones, Zeng, Sidorenko *et al*. (2019), excluding 3,638 SNPs in the MHC region (Chr6 : 28-34Mb) to improve model convergence.

We ran SBayesR assuming a four-component normal mixture model, with initial mixture probabilities *π* = (0.95, 0.02, 0.02, 0.01), and *γ* = (0.0, 0.01, 0.1, 1), where *γ* is a parameter that constrains the variance of the distribution of genetic effects for each of the four components. The MCMC was run for 10,000 iterations with 2,000 taken as burn-in. We obtain the adjusted weights for the overlapping SNPs between the *∼* 2.8 million SNPs available in the LD matrix and the GWAS educational achievement. Using these weights, we calculated the PGS in Plink2 (Chang, Chow, Tellier *et al*., 2015) by multiplying the genotype at each SNP by the corresponding estimated posterior mean calculated by SBayesR, and then summing all included SNPs.

In order to control for population stratification in models that include the PGS, we constructed 20 principal components (PCs) of the genomic data. Prior to constructing the PCs, we excluded SNPs with an imputation accuracy less than 70% or minor allele frequency less than 1%. Next, we obtained a set of approximately independent SNPs by LD-pruning the remaining variants using a 1Mb rolling window (incremented in steps of 5 variants) and an *r*^2^ cutoff of 0.1. We obtained a genetic relatedness matrix using these approximately independent SNPs, and identified all closely related individuals (pairwise relatedness coefficient greater than 0.05 as calculated by Plink 1.9; Chang, Chow, Tellier *et al*. (2015)). We excluded one individual from each of these pairs, estimated the PC loadings in the unrelated sample, and projected the remaining individuals onto the PC space.

## A.5 Imputation

Figure A1 shows the imputation performed on the University of Michigan server via IBS/IBD distance analysis using AIMS variants with a 1000G panel which has the optimal mix (it is the same used for the MCS).

**Figure A1.**
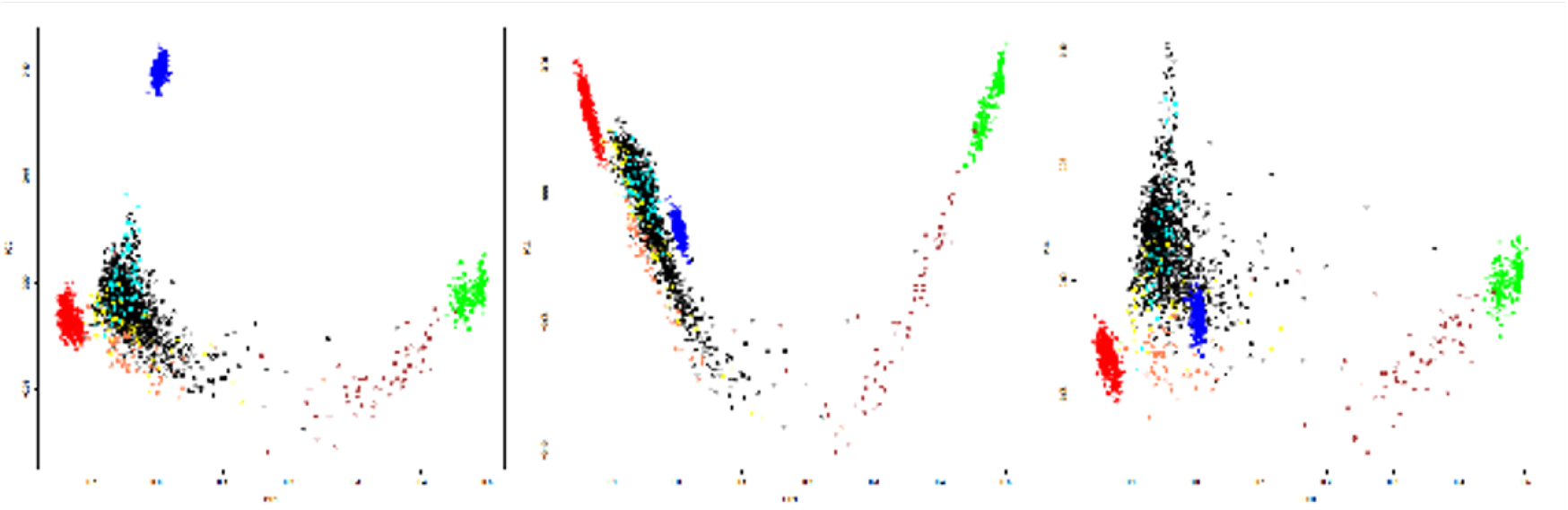
Imputation Note: Our sample is black. Red=European. Blue=Asian. Green=African. Yellow=Colombian. Cyan=Mexican Coral=Puerto Rican. Brown=African American.

## A.6 Balance Test

**Table A1.** Balance.

|  | PC Years of Education | PC Raven score | PC Age | Wealth Index | Play materials | Play activities | PC EA-PGS | Child age | Cognition | Expressive Language | Receptive Language | Child EA-PGS | Bayley Factor |
| --- | --- | --- | --- | --- | --- | --- | --- | --- | --- | --- | --- | --- | --- |
| **Panel A: Child genetic sample** |  |  |  |  |  |  |  |  |  |  |  |  |  |
| Treat | -0.561 | -0.007 | 1.014 | 0.086 | 0.002 | -0.178 | -0.195 | -0.205 | -0.048 | 0.007 | -0.135 | -0.007 | -0.081 |
|  | (0.356) [0.192] | (0.839) [1.000] | (0.572) [0.079] | (0.097) [0.377] | (0.148) [0.128] | (0.170) [0.464] | (0.147) [0.189] | (0.376) [0.587] | (0.097) [0.914] | (0.102) [0.946] | (0.105) [0.412] | (0.137) [0.960] | (0.120) [0.499] |
| Obs. | 493 | 493 | 497 | 511 | 511 | 511 | 334 | 511 | 508 | 507 | 508 | 511 | 507 |
| **Panel B: Child genetic sample, with interviewer fixed effects** |  |  |  |  |  |  |  |  |  |  |  |  |  |
| Treat | -0.670 | -0.052 | 1.047 | 0.077 | 0.034 | -0.120 | -0.323 | -0.080 | -0.081 | -0.024 | -0.198 | -0.051 | -0.139 |
|  | (0.338) [0.081] | (0.864) [0.997] | (0.508) [0.042] | (0.095) [0.420] | (0.157) [0.963] | (0.183) [0.747] | (0.130) [0.015] | (0.387) [0.837] | (0.098) [0.748] | (0.107) [0.828] | (0.096) [0.098] | (0.118) [0.663] | (0.108) [0.204] |
| Obs. | 493 | 493 | 497 | 511 | 511 | 511 | 334 | 511 | 508 | 507 | 508 | 511 | 507 |
| **Panel C: Primary Caregiver genetic sample** |  |  |  |  |  |  |  |  |  |  |  |  |  |
| Treat | -0.254 | 0.190 | 1.354 | 0.117 | 0.017 | -0.297 | -0.195 | -0.038 | -0.178 | -0.060 | -0.211 | 0.009 | -0.193 |
|  | (0.356) [0.766] | (0.839) [0.973] | (0.696) [0.055] | (0.114) [0.309] | (0.148) [0.996] | (0.170) [0.221] | (0.147) [0.189] | (0.477) [0.937] | (0.097) [0.398] | (0.102) [0.642] | (0.105) [0.322] | (0.142) [0.948] | (0.126) [0.129] |
| Obs. | 322 | 323 | 324 | 334 | 334 | 334 | 334 | 334 | 332 | 331 | 332 | 334 | 331 |
| **Panel D: Primary Caregiver genetic sample, with interviewer fixed effects** |  |  |  |  |  |  |  |  |  |  |  |  |  |
| Treat | -0.601 | -0.102 | 1.387 | 0.098 | 0.047 | -0.219 | -0.323 | 0.130 | -0.249 | -0.115 | -0.308 | -0.093 | -0.293 |
|  | (0.338) [0.128] | (0.864) [0.988] | (0.593) [0.022] | (0.115) [0.395] | (0.157) [0.953] | (0.183) [0.448] | (0.130) [0.015] | (0.501) [0.796] | (0.098) [0.272] | (0.107) [0.436] | (0.096) [0.091] | (0.129) [0.473] | (0.122) [0.019] |
| Obs. | 322 | 323 | 324 | 334 | 334 | 334 | 334 | 334 | 332 | 331 | 332 | 334 | 331 |

Note: Table displays results from balancing tests. Panels A and C display coefficients from linear regressions of the baseline characteristics indicated in the column and a binary indicator for treatment. Panels B and D display coefficients from specifications which include binary indicators for interviewer. Clustered standard errors at the village level in parenthesis, and Romano-Wolf adjusted two tailed p-values (Romano and Wolf (2005, 2016)) in squared brackets (5000 replications). For those cases where there is no Romano-Wolf adjustment the p-value is presented in the squared brackets. Romano-Wolf implemented on the following blocks: (1) PC Years of Education and PC Raven Score; (2) Play Materials and Play Activities; (3) Bayley Cognitive, Bayley Expressive Language and Bayley Receptive Language. \* $p < 0.10$ , \*\* $p < 0.05$ , \*\*\* $p < 0.01$ .

## Footnotes

1 As we discuss in Section 2.3, a PGS combines the predictive ability of a large number of Single Nucleotide Polymorphisms (SNPs). These are the locations in the genome where most of the variation in humans is expressed. Each SNP is weighted by the size of the GWAS regression coefficient on an outcome of interest, which is estimated from very large samples. The weighted SNPs are cumulated to compute an aggregate score that represents an individual’s genetic propensity toward a certain outcome.

2 In addition, PGSs have been found to predict a range of other outcomes such as resilience to external events (Amstadter, Moscati, Maes *et al*., 2014), depression (Coleman, Wray, and Lewis, 2020; Lahti, Silventoinen, and Jokela, 2024), and obesity (Khera, Chaffin, Wade *et al*., 2019).

3 Specifically, these authors find that the IGE coefficient may be larger than previously estimated and is influenced by the distribution of genetic endowments within the population.

4 See Conti and Heckman (2010) for one of the first papers laying out a framework for the analysis of gene-environment interactions.

5 These are step-down p-values allowing for testing of 12 hypotheses (Romano and Wolf, 2005).

6 The principal caregiver is the mother in the majority of instances, so from now on we use these terms interchangeably.

7 Here we use this composite factor to increase the power of the analysis. The results for the raw scores for each sub-scale and other domains in the Bayley are reported in Attanasio, Fernández, Fitzsimons *et al*. (2014)

8 We lost a variable proportion of observations within each community (cluster) and only in the two cases we lost the entire cluster. In our data, the spatial correlation is about 4%, implying that the power lost from missing observations is lower than it would have been if the observations had been independent of each other.

9 Tables using the specification with continuous PGS yield very similar results and are available upon request.

10 The alternative specifications with a continuous PGS deliver very similar results, and are presented in the appendix.

11 For those for whom we have DNA, most of whom are the children’s biological mothers.

12 It would have been desirable to also control for the PGS of the principal caregiver, but the number of observations where both are available is too small, so the estimates become very noisy.

13 To ensure proper sample preservation, the staff were trained to seal each tube securely with Parafilm, place them inside pressure-resistant boxes to prevent damage, store them at room temperature in a closed container, track collection progress, and report to the field supervisor. Once collected, the samples were shipped to the Human Population Lab at University College London’s Department of Genetics, Evolution, and Environment, where DNA was stored.

